# Assessing the impact of a mandatory calorie labelling policy in out-of-home food outlets in England on consumer behaviour: a natural experimental study

**DOI:** 10.1101/2024.06.07.24308607

**Authors:** Michael Essman, Thomas Burgoine, Andrew Jones, Megan Polden, Eric Robinson, Gary Sacks, Stephen J. Sharp, Richard Smith, Lana Vanderlee, Christine M. White, Martin White, David Hammond, Jean Adams

**Affiliations:** MRC Epidemiology Unit, University of Cambridge; School of Psychology, Liverpool John Moore’s University; Institute of Population Health Sciences, University of Liverpool; Institute for Health Transformation, Deakin University; Department of Public Health and Sports Science, Faculty of Health and Life Sciences, University of Exeter; School of Nutrition, Centre Nutrition, santé et société (NUTRISS), INAF, Université Laval, Québec, Canada; School of Public Health Sciences, University of Waterloo, Waterloo, Canada

## Abstract

**Background:** Out-of-home (OOH) food tends to be energy-dense and nutrient-poor. In response, England implemented a mandatory calorie labelling policy in the OOH sector. We evaluated changes in consumer behaviours after the policy was implemented in April 2022.

**Methods:** We employed a natural experimental design to assess pre-post changes in noticing and using nutrition information, and behaviours associated with menu labelling. We compared changes in England to comparator jurisdictions without similar policies. Data included four consecutive years (2019-2022) from the International Food Policy Study; participants were adults aged 18 years or older. Mixed effects logistic regression models assessed pre-post changes in binary outcomes, and mixed effects negative binomial regression assessed changes in frequency of OOH eating.

**Results:** In England, noticing nutrition information increased from 16.0% (15.6 to 16.4) in 2020 to 19.7% (19.1 to 20.2) in 2021 and to 25.8% (25.5 to 26.1) in 2022. This increase was 4.8 percentage points (95% CI 2.5 to 7.1) higher in England versus the comparator group. Using nutrition information increased in England from 8.0% (7.5% to 8.4%) in 2020 to 11.8% (10.9% to 12.6%) in 2021 and to 13.5% (13.1% to 13.9%) in 2022. There was a 2.7 percentage point (95% CI 2.0 to 3.4) greater increase in England versus the comparator group from 2020 to 2021. Ordering something different was the only behaviour associated with menu labelling that increased after the policy in England: from 12.6% (12.4 to 12.7) in 2020 to 15.2% (14.7 to 15.6) in 2021 and to 17.7% (17.6 to 17.8) in 2022. There was a 2.8 percentage points (95% CI 1.8 to 3.9) greater increase in England versus the comparator group from 2021 to 2022. Frequency of OOH eating did not change after policy implementation.

**Conclusions:** The introduction of mandatory calorie labelling in England led to increases in self-reported noticing and using, with the key behavioural impact on ordering something different. This suggests that while calorie labelling can enhance awareness, translating this into behaviour change remains limited to shifting orders. Additional strategies may be required to maximize the public health benefits of calorie labelling.

## Introduction

The out-of-home (OOH) food sector includes physical and online locations where food and beverages is sold for immediate consumption including, restaurants, cafés, pubs and bars, takeaways, fast food, street-food and other sites (1). OOH eating has become common in many countries and is increasing globally (1,2), typically involving energy-dense, nutrient-poor foods that contribute to elevated energy intake and increased risk of obesity (3–5). There is also evidence that individuals underestimate the calorie content of foods when eating OOH (6,7), and a recent study in England found that customers of OOH food outlets underestimated calories purchased by an average of 253 kcal (8). In response to the public health challenge posed by the OOH food environment, some governments have adopted policies requiring mandatory calorie labels in the OOH sector to help the public make informed food choices in these settings (9).

A mandatory calorie labelling policy for the OOH food sector was implemented in England on the 6th April 2022. Food businesses are in scope of the policy if they sell food for immediate consumption that is not pre-packaged, and the business has at least 250 employees (10). Exempt establishments include education institutions for pupils <18 years, workplace canteens solely used by employees, and health and social care settings where food is solely provided for patients or residents. Specific item exemptions include menu items available for less than 30 days, beverages with greater than 1.2% alcohol content by volume, unpackaged and unprepared fruit and vegetables, and condiments added by consumers (i.e. not pre-prepared) (10). Calorie labels must display the energy content (kcal) of food for the given portion size and must be accompanied by the reference statement ‘adults need around 2000 kcal a day’ (10). Labels must be easily visible and clearly legible for both online and in-store purchases at all points of choice, defined as any place where customers choose what food to buy (10).

The evidence for the impact of calorie labelling on consumer choices is mixed. A meta-analysis of non-experimental field data found calorie labelling interventions were associated with 21 fewer kcal selected by customers (11). Another meta-analysis of randomized controlled trials found a reduction of 47 kcal purchased after energy labelling was implemented on menus in restaurants (12). Studies in the United States of America (USA) have found small-to-moderate decreases in energy purchased from supermarkets and fast-food restaurants (13,14). However, many studies in real world settings on the effects of calorie labelling policies lack a comparison group (13–15), and those that do are small-scale (16–18). A study in Canada found that mandatory calorie information on menus was associated with greater noticing of nutrition information and that it influenced purchases (19). Overall, there is limited evidence available for the effectiveness of national level calorie labelling policies.

There is also a need to understand the potential mechanisms through which calorie labelling policies may affect consumer choices beyond calories purchased and consumed. The present study examines other consumer behavioural outcomes to inform a better mechanistic understanding of how calorie labelling policies are associated with consumer behaviours at restaurants. A conceptual framework presented by Burton and Kees (2012) describes how calorie labels may affect behaviour. First, customers must be aware of the calorie information; second, they must be motivated to seek healthier items; third, to make the healthier selection they must have knowledge of their daily caloric requirements; and fourth, calorie labelling must provide customers with new information that translates to a different choice than they would have made without labels (20). Consistent with this, policy impact may be limited if customers do not notice labels at sufficient rate to translate to downstream behavioural changes (21). There are also concerns from the food industry that calorie labelling policies could potentially reduce customer patronage if it negatively affects the OOH eating experience, and these potential changes could economically harm the OOH food sector.

The present study is the first study examining pre-post changes in OOH consumer behaviour after implementation of a national calorie labelling policy in a natural experimental framework (22). The aims of this study were to assess whether implementation of the mandatory calorie labelling policy in large OOH food outlets in England was associated with changes in: (1) noticing nutrition information, (2) using nutrition information, and (3) behaviours potentially associated with using nutrition information labels. Changes in England were compared to control jurisdictions without a comparable labelling policy.

## Methods

### Dataset

This study utilised data from Australia, Canada, Mexico, the United Kingdom (UK), obtained from the International Food Policy Study (IFPS), an annual multi-country repeated cross-sectional survey. Designed as a natural experimental framework, the IFPS allows for the evaluation of large-scale food policies within participating countries and facilitates comparisons with included jurisdictions that have not adopted such policies (22). The current analysis included four consecutive years of data (2019-2022) from the IFPS (22).

The study sample for IFPS was recruited from Nielsen Consumer Insights Global Panel and their partners’ panels. A random sample of participants aged 18–100 years were invited to complete the IFPS survey (23). Online surveys were completed between November and December annually. Thus, the post-policy surveys in England were conducted 7-8 months after the April 2022 implementation date.

The conceptual framework used in this study assumes that for nutrition labelling to positively influence eating decisions, nutrition information must be noticed and then used in different ways to promote healthier eating. In addition to noticing and using nutrition information, outcomes included frequency of OOH eating and four other behaviours related to menu labelling at restaurants. Outcomes and potential confounders are defined in Table 1. Participants who visited a restaurant within the last 6 months (Table 1) were asked a series of questions about their behaviours at restaurants, and as such, analyses for all study outcomes were restricted to that population. Sex was chosen as a potential confounder instead of gender identity due to small sample sizes in gender categories other than Male or Female. The IFPS was reviewed by and received ethics clearance through a University of Waterloo Research Ethics Committee (ORE# 30829). A full description of IFPS methods can be found at https://foodpolicystudy.com/methods/.

**Table 1.**
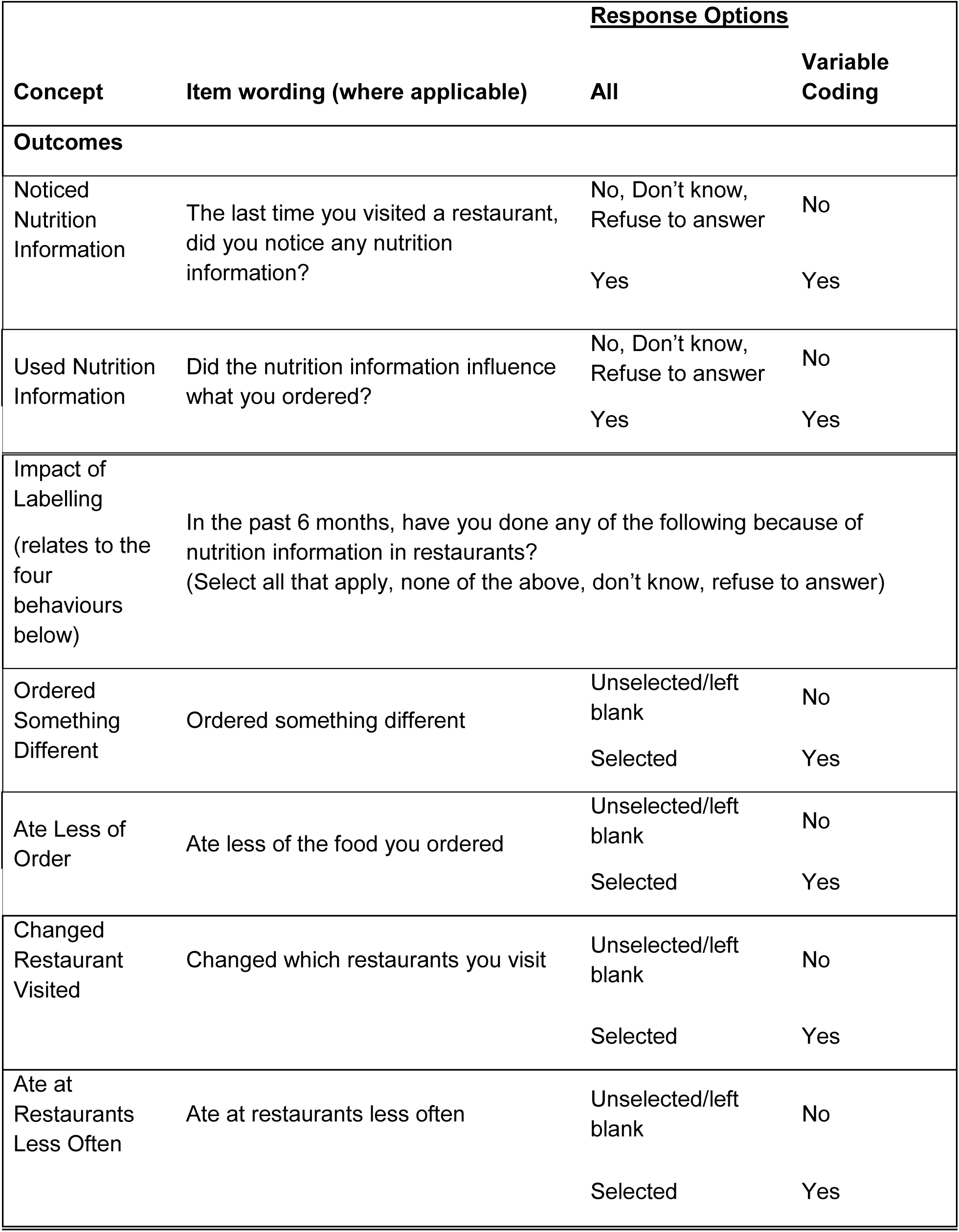
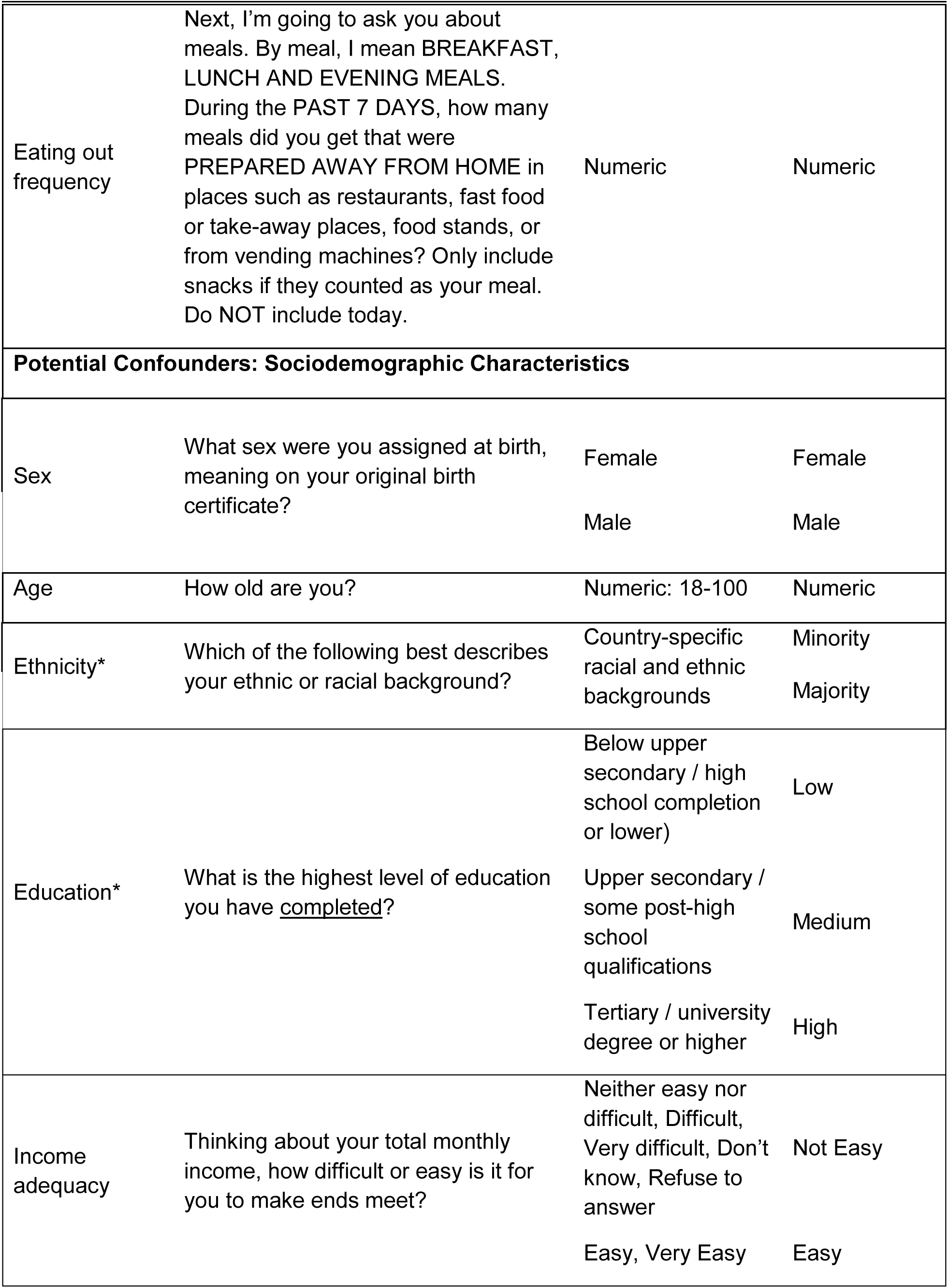
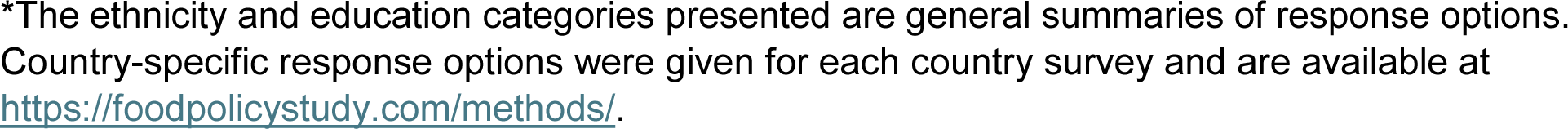
IFPS 2019-2022 survey questions, survey response options, and variable coding for analysis.

|  |  | <u>Response Options</u> |  |
| --- | --- | --- | --- |
| Concept | Item wording (where applicable) | All | Variable Coding |
| Outcomes |  |  |  |
| Noticed Nutrition Information | The last time you visited a restaurant, did you notice any nutrition information? | No, Don't know, Refuse to answer | No |
|  |  | Yes | Yes |
| Used Nutrition Information | Did the nutrition information influence what you ordered? | No, Don't know, Refuse to answer | No |
|  |  | Yes | Yes |
| Impact of Labelling<br>(relates to the four behaviours below) | In the past 6 months, have you done any of the following because of nutrition information in restaurants?<br>(Select all that apply, none of the above, don't know, refuse to answer) |  |  |
| Ordered Something Different | Ordered something different | Unselected/left blank | No |
|  |  | Selected | Yes |
| Ate Less of Order | Ate less of the food you ordered | Unselected/left blank | No |
|  |  | Selected | Yes |
| Changed Restaurant Visited | Changed which restaurants you visit | Unselected/left blank | No |
|  |  | Selected | Yes |
| Ate at Restaurants Less Often | Ate at restaurants less often | Unselected/left blank | No |
|  |  | Selected | Yes |
| Eating out frequency | Next, I'm going to ask you about meals. By meal, I mean BREAKFAST, LUNCH AND EVENING MEALS.<br>During the PAST 7 DAYS, how many meals did you get that were PREPARED AWAY FROM HOME in places such as restaurants, fast food or take-away places, food stands, or from vending machines? Only include snacks if they counted as your meal. Do NOT include today. | Numeric | Numeric |
| **Potential Confounders: Sociodemographic Characteristics** |  |  |  |
| Sex | What sex were you assigned at birth, meaning on your original birth certificate? | Female<br>Male | Female<br>Male |
| Age | How old are you? | Numeric: 18-100 | Numeric |
| Ethnicity* | Which of the following best describes your ethnic or racial background? | Country-specific racial and ethnic backgrounds | Minority<br>Majority |
| Education* | What is the highest level of education you have <u>completed</u>? | Below upper secondary / high school completion or lower)<br>Upper secondary / some post-high school qualifications<br>Tertiary / university degree or higher | Low<br>Medium<br>High |
| Income adequacy | Thinking about your total monthly income, how difficult or easy is it for you to make ends meet? | Neither easy nor difficult, Difficult, Very difficult, Don't know, Refuse to answer<br>Easy, Very Easy | Not Easy<br>Easy |

\*The ethnicity and education categories presented are general summaries of response options. Country-specific response options were given for each country survey and are available at https://foodpolicystudy.com/methods/.

### Study design

The implementation of mandatory calorie labelling in England in 2022 enables examination of behavioural changes across years, comparing England as the intervention country to comparator countries without a policy. The countries within the International Food Policy Study (IFPS) have varying mandatory menu labelling policies, with some mandatory at the national levels, others at the state/province level and others with no mandatory menu labelling policy. Thus, this multi-country survey includes large populations that were and were not exposed to mandatory calorie (or other energy unit) labelling policies at the time of data collection.

This study compared pre-post changes in consumer behaviour in England in 2022 (post-policy) to the pre-policy years of 2019-21. Using a ‘natural experimental’ approach, changes in England from year to year were compared to jurisdictions with no policy for the entire study period. Within this natural experiment framework, England was designated as the intervention, and IFPS jurisdictions with no policy throughout the entire study period were the comparator group. That is: all of Mexico, jurisdictions of Australia and Canada without a policy, and jurisdictions of the United Kingdom without a policy (Table 2).

**Table 2.** Categorization of jurisdictions according to presence or absence of mandatory menu labelling policies before 2019 data collection (9).

| Country and Policy Status | Jurisdiction | Description | Unweighted n for this analysis |
| --- | --- | --- | --- |
| **Intervention group** |  |  |  |
| England | National policy (2022) | In April 2022, England introduced mandatory calorie menu labelling for large out-of-home food businesses with more than 250 employees. | n=11,732 |
| **Comparator group – no policy present** |  |  |  |
| Australia – jurisdictions without a policy | Western Australia, Tasmania, Northern Territory | The three states/territories included in the analysis do not have a mandatory menu labelling policy. Other states/territories with policies were excluded from the analysis. | n=1,719 |
| Canada – jurisdictions without a policy | All provinces other than Ontario | Provinces other than Ontario do not have a mandatory menu labelling policy. Ontario implemented a mandatory menu labelling policy in 2017, and was excluded from the analysis. | n=9,752 |
| Mexico – no policy | Whole country | No mandatory menu labelling policy. | n=14,494 |
| United Kingdom – jurisdictions without a policy | Scotland, Wales, and Northern Ireland | No mandatory menu labelling policy. | n=1,928 |

### Modelling approach

All statistical analyses were conducted in Stata 17 (24). Descriptive statistics were calculated to summarize sociodemographic characteristics of the study sample by year and for England and the comparator group separately. Binary outcomes were the six outcome variables in Table 1. Binary outcomes were modelled using survey-weighted mixed effects logistic regression, with clustering at the country level. Post-stratification sample weights were constructed using a raking algorithm with population estimates in each country separately based on age group, sex, region, and (except in Canada) ethnicity. Weights were subsequently rescaled to each sample size. Models were adjusted for potential confounders listed in Table 1, and included indicator variables for England vs comparator group and study year. To estimate the potential differences in pre-post changes between England and the comparator, two-way interactions between policy group and study year were included. The marginal probability of each outcome was calculated by year and policy status (25). Difference-in-differences were calculated for the changes from each consecutive year (2019 to 2020, 2020 to 2021, 2021 to 2022) in England compared to the changes in those years in the comparator. To explore the potential for spillover effects between England and the rest of the UK, we performed sensitivity analyses using the same outcomes, but we separated the rest of the UK countries (Scotland, Wales, and Northern Ireland) into a third group and described trends and outcomes.

A similar approach was used for the outcome frequency of eating food prepared away from home (online or in-person) in the last seven days using survey-weighted negative binomial regressions. Marginal means were calculated by year and policy status, and difference-in-differences were calculated for the changes from each consecutive year (2019 to 2020, 2020 to 2021, 2021 to 2022) in the policy group compared to the changes in those years in the comparator (25).

## Results

A total of 67,960 adults completed the IFPS surveys for 2019–2022 across the four countries. 46,809 people met the inclusion criteria described in Table 2 of being either from England or a jurisdiction without a comparable menu labelling policy. 40,209 (85.9%) participants reported visiting a restaurant within the last 6 months and answered the questions for the outcomes used for this analysis. Of this sample size that met all inclusion criteria, 467 observations (1.2%) were removed due to missing data on ethnicity, and a further 117 observations (0.3%) were removed due to missing data on education. The final sample included 39,625 respondents (2019 = 10,737; 2020 = 8,609; 2021 = 9,967; 2022 = 10,312).

Table 3 describes the sample characteristics, stratified by policy status and year. Most of the sample reported high education and low-income adequacy (i.e. not easy to make ends meet). The distribution of education was similar between England and the comparator across years, although there were more low education participants surveyed from England in 2020. Participants in the comparator were slightly older than in England.

**Table 3.** Sample characteristics (data are unweighted n, weighted %; or weighted mean (SD)

|  | **2019**<br>**(pre-implementation)** |  | **2020**<br>**(pre-implementation)** |  | **2021**<br>**(pre-implementation)** |  | **2022**<br>**(post-implementation)** |  |
| --- | --- | --- | --- | --- | --- | --- | --- | --- |
|  | **England** | **Comparator** | **England** | **Comparator** | **England** | **Comparator** | **England** | **Comparator** |
| **Variable** | **n=3194**<br>n, % | **n=7543**<br>n, % | **n=2489**<br>n, % | **n=6120**<br>n, % | **n=2906**<br>n, % | **n=7061**<br>n, % | **n=3143**<br>n, % | **n=7169**<br>n, % |
| **Sex** |  |  |  |  |  |  |  |  |
| Male | 1574, 49.0 | 3761, 48.6 | 1249, 49.7 | 3130, 50.0 | 1446, 48.5 | 3534, 49.4 | 1528, 48.2 | 3520, 48.8 |
| Female | 1620, 51.0 | 3782, 51.4 | 1240, 50.3 | 2990, 50.0 | 1460, 51.5 | 3527, 50.6 | 1615, 51.8 | 3649, 51.2 |
| **Ethnicity** |  |  |  |  |  |  |  |  |
| Majority | 2865, 87.4 | 6360, 81.5 | 2167, 85.2 | 5133, 81.5 | 2528, 86.6 | 5892, 82.0 | 2706, 83.2 | 5952, 81.4 |
| Minority | 329, 12.6 | 1183, 18.5 | 322, 14.8 | 987, 18.5 | 378, 13.4 | 1169, 18.0 | 437, 16.8 | 1217, 18.6 |
| **Income Adequacy** |  |  |  |  |  |  |  |  |
| Not easy | 1739, 59.5 | 5518, 75.4 | 1456, 60.0 | 4436, 74.1 | 1527, 56.8 | 4807, 70.7 | 2015, 66.8 | 5291, 75.3 |
| Easy | 1455, 40.5 | 2025, 24.6 | 1033, 40.0 | 1684, 25.9 | 1379, 43.2 | 2254, 29.3 | 1128, 33.2 | 1878, 24.7 |
| **Education** |  |  |  |  |  |  |  |  |
| Low | 914, 49.8 | 1875, 31.9 | 933, 48.6 | 1630, 32.6 | 838, 47.1 | 1595, 31.0 | 836, 37.7 | 1562, 29.5 |
| Medium | 821, 20.9 | 1846, 21.9 | 742, 20.3 | 1627, 23.1 | 798, 22.6 | 1805, 22.9 | 798, 25.6 | 1828, 22.0 |
| High | 1459, 29.4 | 3822, 46.2 | 814, 31.1 | 2863, 44.4 | 1270, 30.3 | 3661, 46.1 | 1509, 36.7 | 3779, 48.5 |
| **Age** |  |  |  |  |  |  |  |  |
| Mean (SD) | 48.0 (16.9) | 43.2 (15.9) | 45.5 (17.2) | 43.1 (16.0) | 47.5 (17.3) | 44.2 (16.3) | 47.3 (17.3) | 44.1 (16.2) |

### Noticed nutrition information

There were no significant differences in noticing nutrition information between years in the comparator. In England, the probability of noticing nutrition information increased from 16.0% (15.6 to 16.4) in 2020 to 19.7% (19.1 to 20.2) in 2021 with a further increase to 25.8% (25.5 to 26.1) in 2022 (Fig. 1a). There was no evidence of a difference in changes between England and the comparator in 2020 vs 2019. For 2021 vs 2020, the difference in the probability of noticing was 2.9 percentage points (95% CI 1.7 to 4.1) higher in England compared to the comparator. For 2022 vs 2021, the difference was 4.8 percentage points (95% CI 2.5 to 7.1) higher in England compared to the comparator (Fig. 3).

**Figure 1a-b.**
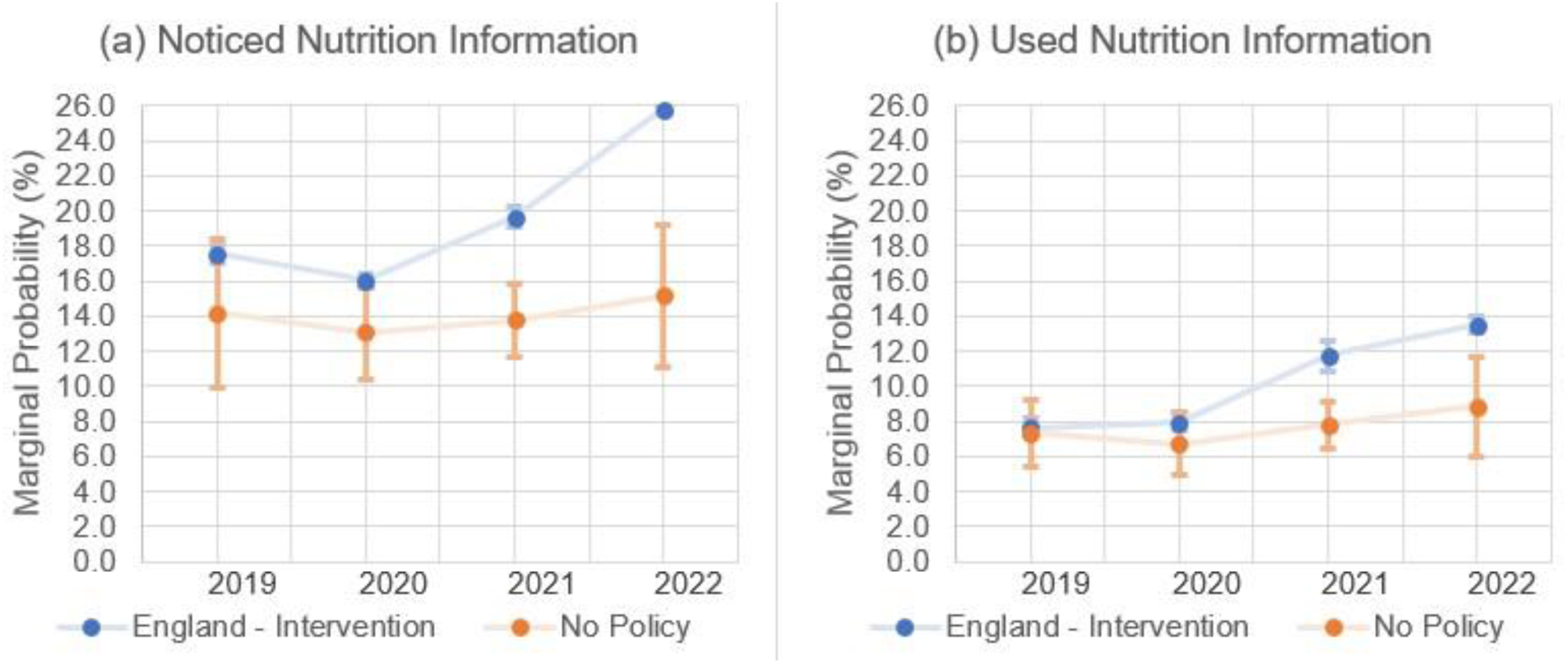
Marginal probability of (a) noticing and (b) using nutrition information from 2019-2022 for England and the comparator estimated from mixed effects logistic regression model adjusted for age, sex, education, perceived income adequacy, and ethnicity. Error bars represent 95% confidence intervals and are presented for both England and the comparator. The confidence intervals for England are narrow, at ±0.6% or less for the outcomes.

### Used nutrition information

In England, the probability of using nutrition information increased from 8.0% (7.5% to 8.4%) in 2020 to 11.8% (10.9% to 12.6%) in 2021 and further increased to 13.5% (13.1% to 13.9%) in 2022 (Fig. 1b). There was no evidence of a difference in changes between England and the comparator in 2020 vs 2019. For 2021 vs 2020, the difference in the probability of using nutrition information was 2.7 percentage points (95% CI 2.0 to 3.4) higher in England compared to the comparator. For 2022 vs 2021, the difference was smaller and not statistically significant (Fig. 3).

### Ordered Something Different

There were no significant differences in ordering something different because of nutrition labelling between years in the comparator. There was a slight reduction in ordering something different in 2020 vs 2019 in England, after which there were significant increases in England in 2021 and 2022 (Fig. 2a). In England, the probability of ordering something different increased from 12.6% (12.4 to 12.7) in 2020 to 15.2% (14.7 to 15.6) in 2021 and a further increase to 17.7% (17.6 to 17.8) in 2022 (Fig. 2a). For 2022 vs 2021, the difference in the probability of ordering something different was 2.8 percentage points (95% CI 1.8 to 3.9) greater in England compared to the comparator (Fig. 3).

**Figure 2a-2d.**
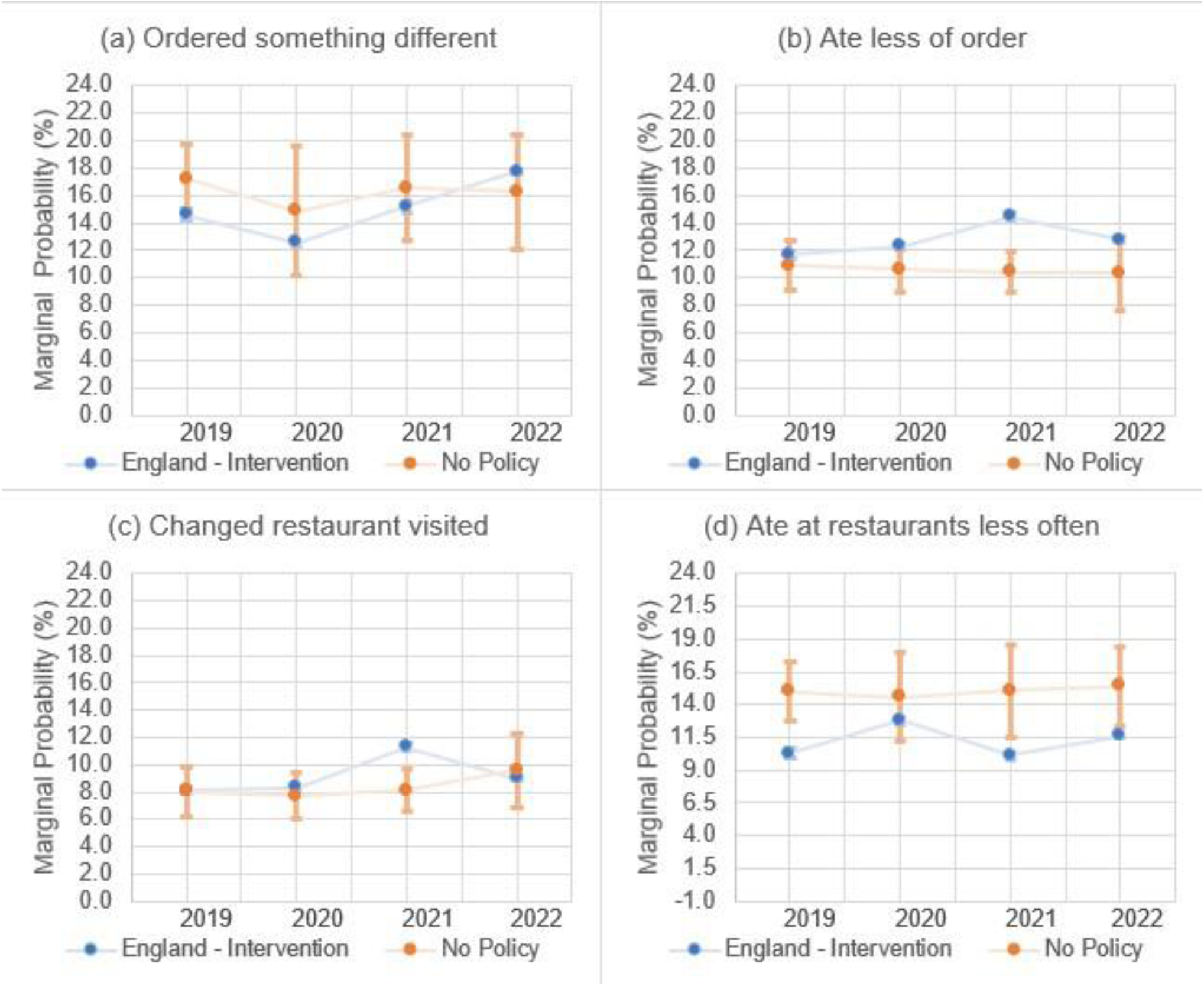
Marginal probability (%) of (a) ordered something different, (b) ate less of order, (c) changed restaurants visited, and (d) ate at restaurants less often from 2019-2022 for England and the comparator. Estimations from mixed effects logistic regression model adjusted for age, sex, education, perceived income adequacy, and ethnicity. Error bars represent 95% confidence intervals and are presented for both England and the comparator. The confidence intervals for England are narrow, at ±0.5% or less for the outcomes.

**Figure 3.**
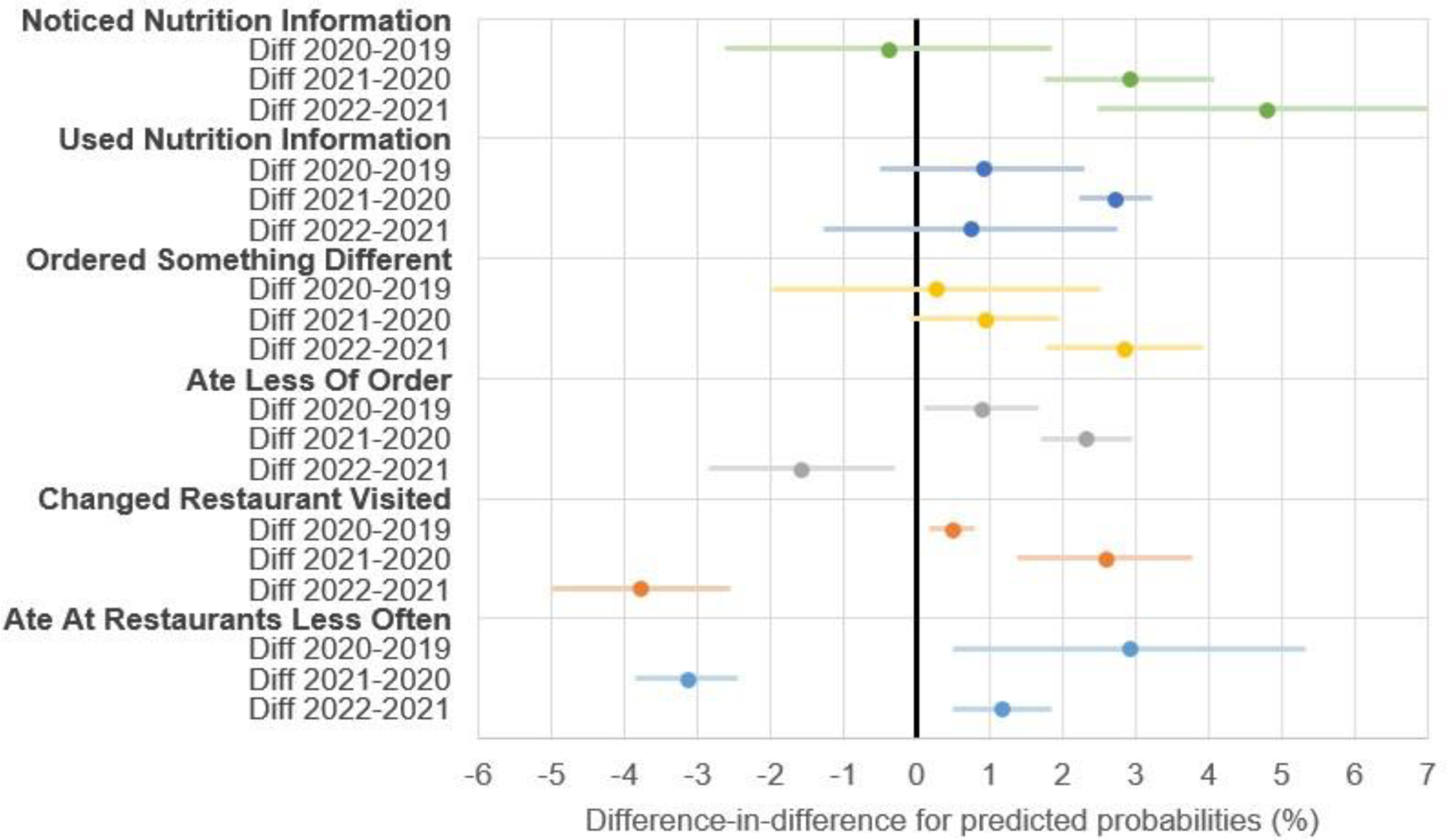
Difference-in-differences between years for each behavioural outcome estimated from mixed effects logistic regression model adjusted for age, sex, education, perceived income adequacy, and ethnicity.

### Ate less of food ordered

There were no significant differences in eating less of the food ordered because of nutrition labelling between years in the comparator. In England, the probability of eating less of the food ordered increased from 12.3% (12.1 to 12.5) in 2020 to 14.4% (14.2 to 14.7) in 2021 and reduced to 12.8% (12.6 to 13.0) in 2022 (Fig. 2b). There was no evidence of a difference in changes between England and the comparator in 2020 vs 2019. For 2021 vs 2020, the difference in the probability of eating less of the food ordered was 2.3 percentage points (95% CI 1.7 to 2.9) greater in England compared to the comparator (Fig. 3). For 2022 vs 2021, the difference in the probability of eating less of the food ordered was 1.6 percentage points (95% CI 0.3 to 2.9) lower in England compared to the comparator.

### Changed restaurants visited

There were no significant differences in changing restaurants visited because of nutrition labelling between years in the comparator. In England, the probability of changing restaurants visited increased from 8.3% (95% CI 8.2 to 8.4) in 2020 to 11.3% (95% CI 11.0 to 11.6) in 2021 and reduced to 9.0% (95% CI 8.8 to 9.2) in 2022 (Fig. 2c). For 2020 vs 2019, the change in the probability of changing restaurants visited was 0.5 percentage points (95% CI 0.2 to 0.8) greater in England compared to the comparator (Fig. 3). In 2021 vs 2020, the difference in the probability of changing restaurants visited was 2.6 percentage points (95% CI 1.4 to 3.8) greater in England compared to the comparator (Fig. 3). In 2022 vs 2021, the difference in the probability of changing restaurants visited was 3.8 percentage points (95% CI 2.6 to 5.0) lower in England compared to the comparator.

### Ate at restaurants less often

There were no significant differences in eating at restaurants less often because of nutrition labelling between years in the comparator. The probability of eating at restaurants less often was lower in England compared to the comparator in all years (Fig. 2d). For 2020 vs 2019, the difference in the probability of eating at restaurants less often was 2.9 percentage points (95% CI 0.5 to 5.3) greater in England compared to the comparator (Fig. 3). For 2021 vs 2020, the difference in the probability of eating at restaurants less often was 3.1 percentage points (95% CI 2.4 to 3.8) lower in England compared to the comparator, as England returned to baseline levels after 2020 (Fig. 2d; Fig. 3). For 2022 vs 2021, the difference in the probability of eating at restaurants less often was 1.2 percentage points (95% CI 0.5 to 1.8) greater in England compared to the comparator (Fig. 3).

### Frequency of eating out

Frequency of eating out decreased from 2019 to 2020 in both England and the comparator (Fig. 4a). For the difference-in-difference results, there was no significant differences between years comparing changes in England to the comparator.

**Figure 4a-4b.**
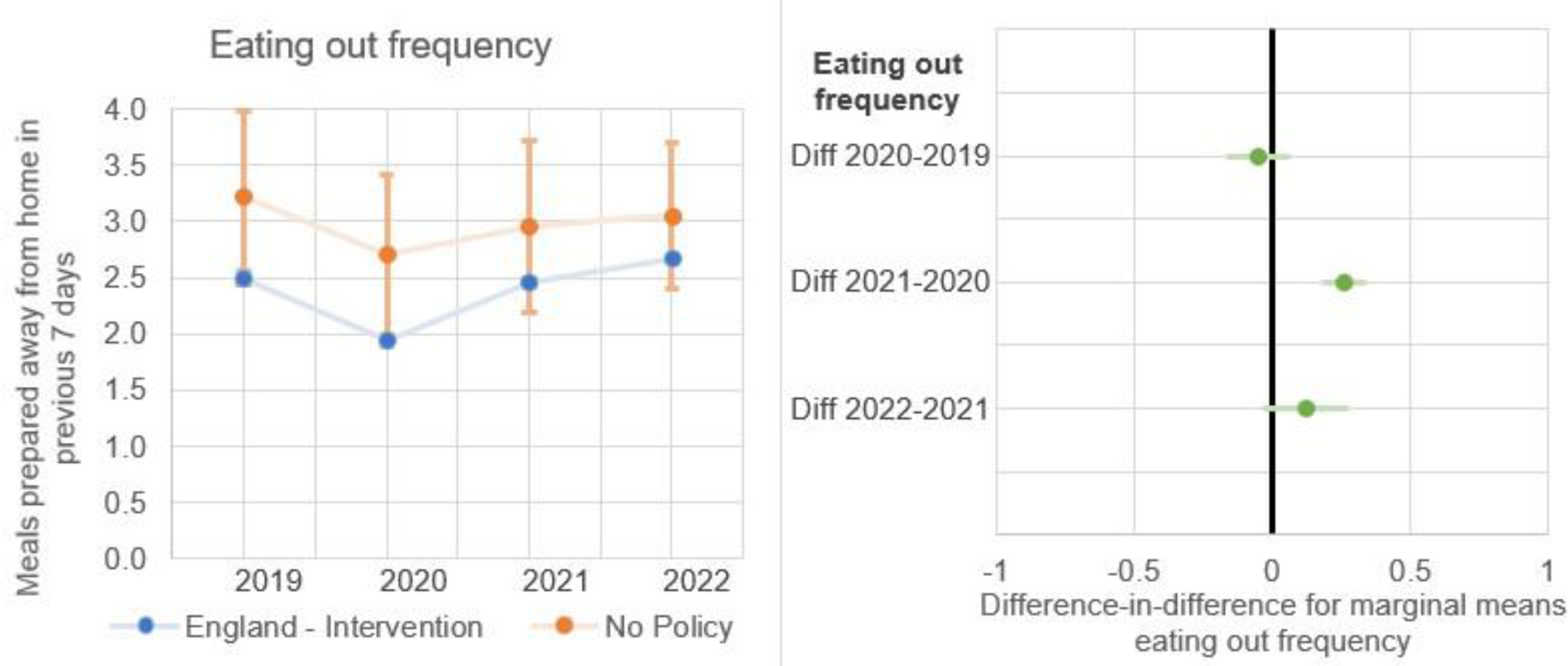
(a) Frequency of eating out of home from 2019-2022 for England and the comparator, (b) Difference-in-differences between years for frequency eating out of home.

### Sensitivity analysis

The sensitivity analysis found substantial evidence of spillover effects from England to the rest of the UK. Similar to England, the rates of noticing and using nutrition information in the rest of the UK increased from 2021 to 2022 (Supplementary Fig. 1a, b). Rates of ordering something different in the rest of the UK also closely tracked those in England (Supplementary Fig. 1c). Although rates of eating less of the food ordered were lower in the rest of the UK in 2020 and 2021, they increased to a similar level as England in 2022 (Supplementary Fig. 1d). Rates of changed restaurant visited in the rest of the UK closely tracked those in England (Supplementary Fig. 1e). Rates of eating at restaurants less often in the rest of the UK followed similar trends to England from 2019-2022 (Supplementary Fig. 1f). Given the rest of the UK was included in the comparator group in the main analysis, these spillover effects likely diluted any policy effects identified in the main analysis.

## Discussion

### Statement of principal findings

We observed an increase in self-reported noticing and using nutrition information in England after the mandatory calorie labelling policy, and these increases were larger than comparator jurisdictions without a comparable policy. However, when examining how labelling was used, the only consistent change compared to comparator jurisdictions was an increase in ordering something different. We did not find evidence of participants eating OOH less often after policy implementation in England. Some behavioural changes occurred in England in 2021, which may have been due to restaurants implementing calorie labelling in preparation for the official policy implementation date in April 2022.

### Strengths and limitations of the study

This is the first multi-country study to examine changes in behaviours associated with menu labelling after implementation in the OOH food sector, allowing comparisons to jurisdictions without policy implementation. This approach enhances the robustness of our analysis of national-level policies compared to relying on pre-post assessments without comparators. While the comparator group included varying contexts that may influence behaviours related to menu labelling, grouping jurisdictions without a labelling policy together helped mitigate any country-specific influences, enhancing generalisability. The large study population increases statistical power and the ability to observe small effect sizes that could nevertheless have public health impact. There was high internal consistency with the same questions asked across time and place (22). The inclusion of multiple years prior to the policy implementation in England in 2022 (2019-2021), serves as a more robust baseline than a typical before-and-after study using single data points before and after. This extended timeframe, including pre-COVID data from 2019, offers additional context for interpreting changes potentially influenced by the COVID-19 pandemic in 2020 and 2021. Additionally, our study’s diversity of outcomes explores the association between implementation of the policy in England and mechanisms of policy effect on consumer behaviour, beginning with noticing menu labels followed by types of use.

Our study has limitations. The reliance on self-reported behaviours introduces bias inherent in surveys, but we assume this bias was consistent across survey years and countries, limiting the impact on our assessment of change. While trends in comparator jurisdictions remained relatively stable from 2019 to 2022, the specific impact of the intervention may be specific to England. We included the rest of the UK in the comparator group, but found some evidence of spillover effects from England to the rest of the UK. If anything, this would diminish our ability to detect a difference between England and the comparator. Similar spillover effects could potentially exist in Canada and Australia, where some jurisdictions have labelling laws and other areas do not. However, the existence of spillover would suggest that our estimates of changes in England are conservative. Causality remains inferential despite the controlled before-and-after study design, with the possibility of unaccounted-for co-interventions influencing our outcomes of interest. We saw some reductions in OOH eating frequency in 2020, likely due to Coronavirus disease (COVID-19) related business closures, but frequency returned to baseline levels in 2021. We expect that COVID-19 related impacts to OOH eating impacted intervention and comparator groups similarly. Additional concerns regarding background trends in each policy group are somewhat mitigated by the inclusion of three baseline years. However, an interrupted time series analysis may better account for pre-existing time trends, as our comparator jurisdictions may not capture all external factors influencing the study outcomes over time. The study’s analysis period covers 2019-2022, approximately seven to eight months post-policy implementation in England and future work could follow up further. The generalisability of this study to other countries could be limited if specific policies differ in a way that is related to consumer responses. For example, both the size and placement of calorie labels can modify the effects of menu labels on calories purchased (26).

### Interpretation and implications of findings

The results of this study align with previous evidence that mandatory calorie labelling policies can increase noticing of nutrition information. In England, pre-post exit-surveys of 6,578 OOH customers found that 16.5% of participants reported noticing calorie labels pre-policy and 31.8% reported noticing calorie labels post-policy. However, the authors also found no evidence of change in the energy content of purchases pre-post policy implementation (27). This consistency in prevalence of noticing nutrition information is particularly notable given the different types of data collected between the two studies. Although the current study found that noticing nutrition information increased after mandatory calorie labelling in England, there is still room for improvement as rates of noticing remained below 30%. Rates of noticing in the UK are less than those reported in some studies based in the United States, which could be due to labelling prominence. Previous research in England found that about two-thirds of businesses sampled had clear or legible calorie information, and only 15% followed all compliance criteria (28). There may also be a diminution of effect on the pathway from noticing to using nutrition information. Recent work found only 22% of people in England who noticed nutrition information also reported using it (27). Policymakers who are considering implementing mandatory menu labels may consider how to make labels more noticeable and how to enhance the effects of labels. For example, greater display size, increased use of colour, and consumer familiarity with labels are associated with greater attention to labels (30). Customers eating OOH might also prioritize factors other than health, such as indulgence, financial considerations, or convenience (31). Understanding consumer expectations and the mechanisms through which people interact with the OOH food environment is needed to inform policies that align with real-world behaviours. Greater public communication that increases motivation to change may also improve the effects of the policy (32).

We also found that although the mandatory calorie labelling policy was associated with greater noticing than the comparator, there were smaller effects on using calorie information and ordering something different, suggesting a diminishing effect along the potential chain of effect leading to changes in dietary behaviour. Previous work has also identified this diminution of effect between noticing and using nutrition information. A cross-sectional analysis of noticing and using calorie labels at a fast-food chain in the United States found that 60% of participants noticed calorie labels on menus, but only 16% reported using them (29).

There were some changes in 2020 that were potentially due to the influence of the COVID-19 pandemic on OOH eating behaviours. Ordering something different slightly decreased in 2020, but it increased each subsequent year in England. There was an increase in reporting eating at restaurants less often during 2020 in England, but responses returned to baseline rates in subsequent years. Eating out frequency decreased in 2020 in both England and the comparator group, but both returned to near 2019 levels in subsequent years.

Despite the absence of mandatory labelling policies elsewhere in the UK, sensitivity analyses revealed similar trends to those observed in England, suggesting possible spillover effects. These spillover effects may have occurred if it was more efficient for international companies that do business in the UK to implement the same menu changes across all UK countries.

### Unanswered questions and future research

Future research may benefit from exploring more aspects of OOH eating in response to calorie labelling policies to more clearly understand how it impacts all steps of the putative causal pathway to dietary change. It is also unknown whether there are differential behavioural responses to mandatory calorie labelling policies according to individual or eating occasion characteristics. For example, baseline nutrition knowledge and motivation to change may influence the impact of labelling policies and some eating occasions, such as dining out for special occasions, may be more resistant to change than others. Longer follow-up periods could provide a more thorough understanding of policy impact. There is growing evidence from grocery retail that interpretive labels result in more change in purchasing or greater ease of use than simple quantitative information, although the effects of interpretive labels may also depend on the specific type of label and the context (33–36). Further work is required to understand the impact of interpretive nutrition labels (e.g. traffic light and warning labels) in the OOH sector. Finally, more work is needed to determine whether the increases in noticing nutrition information can translate into behaviour change by identifying and addressing barriers along the putative pathway of causation from labelling to behaviour change (37).

## Conclusions

The introduction of a mandatory calorie labelling policy in England was associated with increases in noticing nutrition information, using nutrition information, and ordering something different and these changes were greater than in control jurisdictions without a policy. There was no evidence that the introduction of the policy was associated with changes in eating less of order, changing restaurant visited, or frequency of eating at restaurants. Further work is required to translate changes in noticing and using menu labels into health promoting behavioural changes.

## Supporting information

STROBE Checklist

## Ethics statement

The International Food Policy Study was reviewed by and received ethics clearance through a University of Waterloo Research Ethics Committee (ORE # 30829).

## Data availability statement

Data are available upon reasonable request to the International Food Policy Study team (see www.foodpolicystudy.com).

## Supplementary Materials

**Supplementary Fig 1a-1f.**
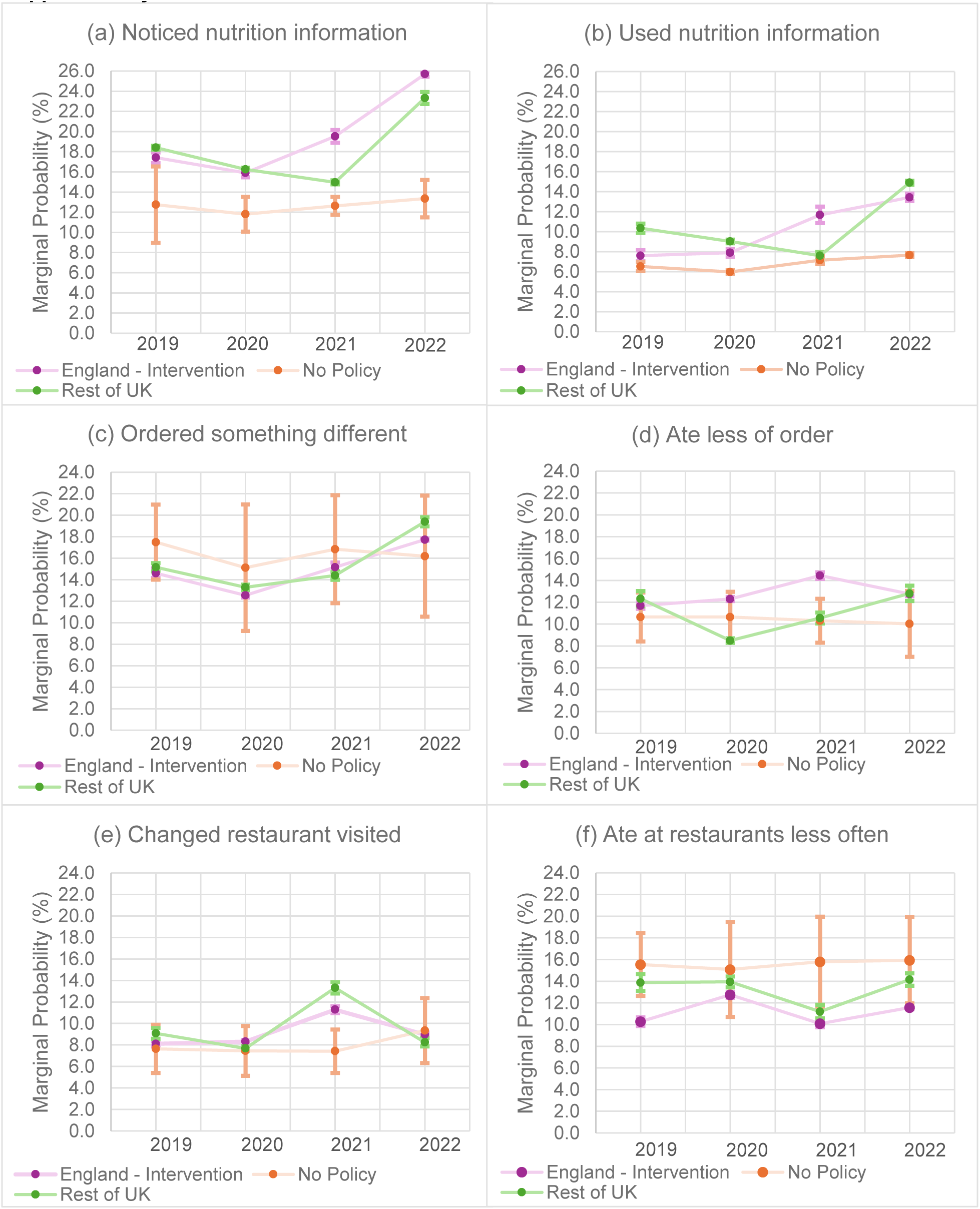
Marginal probability (%) of (a) noticed nutrition information, (b) used nutrition information, (c) ordered something different, (d) ate less of order, (e) changed restaurants visited, and (f) ate at restaurants less often from 2019-2022 for England, No Policy, and Non-England UK.

